# Valvular Heart Disease Associations with Cardiac Biomarkers Using AI-guided Echocardiography: the RURAL Cohort Study

**DOI:** 10.1101/2025.08.27.25334591

**Authors:** Evangelia Alexopoulos, Joanna Walsh, Jesse Y. Hsu, Michael J. Blaha, Susan Cheng, Melissa A. Daubert, Gary Dunn, Peter Durda, Ervin Ray Fox, Yngvil Thomas, Edwin van den Heuvel, Suzanne E. Judd, Ramachandran S. Vasan, Pamela S. Douglas, Gerald S. Bloomfield

## Abstract

**Background:** Few studies have evaluated the prevalence or severity of mitral valve prolapse (MVP) and other valvular heart disease (VHD) in the rural US South, where strategies for early detection are crucial for risk stratification and prevention.

**Objectives:** We assessed the prevalence of MVP and other VHD in a rural US South cohort and examined associations with cardiovascular disease (CVD) risk. We also evaluated relationships between MVP severity, high-sensitivity cardiac troponin T (hsTnT), and N-terminal pro-B-type natriuretic peptide (NTproBNP).

**Methods:** We conducted a cross-sectional analysis from the Risk Underlying Rural Areas Longitudinal (RURAL) study. Logistic regression assessed associations between participant characteristics and MVP, other VHD, or both. Weighted models assessed odds for MVP and other VHD by 10-year CVD risk categories using the Predicting Risk of CVD Events (PREVENT) score. Among a subset, we evaluated associations between MVP severity and cardiac biomarkers.

**Results:** Among 2,621 participants (68.7% women), MVP and other VHD were present in 1.9% and 11.2%, respectively. Compared to the low PREVENT risk group, odds of MVP were lower and odds of VHD were higher among borderline and intermediate/high groups. HsTnT was lower in MVP vs. non-MVP (0.64, 95% CI 0.58-0.71), without difference by severity of MVP. NTproBNP was higher in participants with severe MVP than non-MVP (2.05, 95% CI 1.49-2.83).

**Conclusions:** MVP prevalence aligned with population-based epidemiologic studies. PREVENT risk category may differentiate individuals at higher risk for MVP and for other VHD. Future studies are needed to evaluate relationships between MVP/VHD status and clinical events.

## Introduction

Cardiovascular disease (CVD) disproportionately impacts individuals in rural America, and the rural-urban disparity is widening.^1^ In 2021, Southern US high-poverty rural areas had the highest CVD death rate (256 per 100,000) and the largest disparity relative to low-poverty urban areas (RR: 2.05; 95% CI: 2.01, 2.09).^2^ Socioeconomic risks paired with high rates of co-morbid conditions, such as diabetes and obesity, are amplified in a region with diagnostic barriers and limited specialty care.^3–5^ Addressing these inequalities requires overcoming limited access to the cardiac testing essential for accurate diagnosis.

The epidemiology of CVD and rural-urban disparities has primarily focused on atherosclerotic CVD and data on valvular heart diseases (VHD) are sparse in rural areas.^6,7^ Research has largely addressed acquired VHD in older populations.^8^ Mitral valve prolapse (MVP) is a distinct, morbid subset of VHD that likely affects ∼2% of the population.^9–12^ MVP is the most common cause of mitral regurgitation in western countries and women.^13^ Sudden cardiac death is reported in individuals with MVP with an incidence of 217 events per 100,000 person-years.^14–17^ Despite the higher burden of CVD mortality, rural US regions have historically not been represented in MVP and VHD prevalence estimates. In recognition of this public health threat, the US passed the Cardiovascular Advances in Research and Opportunities Legacy (CAROL) Act in 2023, supporting research into risk factors for cardiac events from VHD, particularly MVP.

Population-based cohort studies have shown morbidity and mortality risks with cardiac biomarker elevations, though the relationship between VHD and biomarker levels is less clear.^18–21^ Elevations in high-sensitivity cardiac troponin (hsTnT) and N-terminal pro-B-type natriuretic peptide (NTproBNP) indicate myocardial injury and abnormal myocardial stretch, respectively. Understanding MVP burden and severity and its association with cardiac biomarkers may yield critical insights for cardiovascular health in rural US communities.

Leveraging the Risk Underlying Rural Areas Longitudinal (RURAL) echocardiography ancillary study we aimed to: (1) assess MVP and other VHD burden in rural southern US counties, (2) identify factors associated with MVP, other VHD, and any VHD, (3) evaluate the relationship between individual CVD risk category and MVP/other VHD, and (4) in a subset, assess the associations between MVP and levels of hsTnT and NTproBNP.

## Methods

### RURAL ECHO study

RURAL ECHO (R01HL157531) is an ancillary study of RURAL (U01HL146382), an NHLBI-funded cohort aiming to characterize individuals in four Southern states with high CVD mortality rates: Alabama, Kentucky, Louisiana, and Mississippi. Using a self-contained mobile examination unit (MEU), RURAL performed detailed baseline examinations including clinical, demographic, non-medical determinants of health, neighborhood, physiologic, biochemical, and multimodality imaging data. Full details of RURAL ECHO have been published^22^; the study was reviewed and approved by IRB (Pro00107822). CAROL Act (NOT-HL-23-079) supplemental funding supported MVP/VHD specific analyses in a subset of participants.

### Study population and sampling

Participants included 2,621 individuals enrolled in RURAL ECHO, selected via a combination of convenience and probability sampling from counties in Mississippi (Panola and Oktibbeha), Alabama (Wilcox), and Louisiana (Assumption and Franklin) as determined by the aims of the CAROL Act supplement (**Central Illustration**). The parent RURAL study includes 10 rural counties—six with higher and four with lower age-adjusted CVD mortality rates than the rest of the country matched within state by poverty level, demographics, and population size using an ecologically paired design.^22^ For MVP and VHD analyses, inclusion was limited to participants with echocardiographic data in counties in Mississippi, Alabama, and Louisiana as the initial enrolling sites in RURAL. For biomarker analyses, a random subset of 1,000 individuals from counties in Alabama and Mississippi with both laboratory and high-quality echocardiography data were included.

### Image Acquisition Using AI-guided Echo

RURAL utilizes FDA-authorized software (Caption Guidance, Caption Health, GE) with deep learning algorithms to guide untrained operators in acquiring transthoracic echocardiograms.^22^ Image acquisition and views are described in **Supplemental Methods**. Echoes were reviewed blinded to race, age, body mass index (BMI), and sex. Participants were categorized as having MVP, VHD, or any VHD (MVP or VHD).

### Echocardiographic Measurements for MVP and VHD

Echocardiograms were analyzed using standardized methods from the American Society of Echocardiography.^23^ MVP was defined as posterior displacement of any portion of the mitral valve leaflets beyond mitral annular plane during ventricular systole of at least 2mm. Other VHD included moderate or greater valvular regurgitation without MVP and other structural abnormalities including leaflet thickening, restriction, calcification, rheumatic changes, bicuspid, or other congenital abnormalities. ‘Any VHD’ refers to MVP, VHD, or both. Severe MVP was defined as MVP plus any of several echocardiographic findings indicating physiologic strain: enlarged LV internal dimension in diastole (>= 5.3 cm in women and >= 5.9 cm in men), LV end diastolic volume >= 87 ml/m2, LV end systolic volume >= 37 ml/m2, LVEF < 50%, left atrial volume >= 34 ml/m2, right ventricular fractional area change < 25%, peak atrial longitudinal strain < 15%, and LV global longitudinal strain > -16%. For severe MVP and other VHD, participants were categorized as not having the disease if at least two findings were non-missing and indicated absence of disease and were categorized as having a missing disease status otherwise. Methods for assessing cardiac structure and function are in the **Supplemental Methods**.

### Clinical Data Definitions

Demographics included sex, age, BMI, and self-reported race. Individual CVD risk was calculated using the American Heart Association’s Predicting Risk of CVD EVENTs (PREVENT) equations for 10-year risk incorporating age, sex, HDL cholesterol, non-HDL cholesterol, systolic blood pressure, diabetes, current smoking, estimated glomerular filtration rate (eGFR), and participant-reported medication use.^24^ Finally, CVD risk was grouped into Low (<5%), Borderline (≥5% to < 7.5%), and Intermediate/High (≥7.5%) per PREVENT guidelines. Cardiac biomarkers assay details are in **Supplemental Methods**.

### Statistical Analyses

Unweighted descriptive statistics are reported for clinical and demographic variables: medians and IQRs are reported for numerical variables and percentages are reported for categorical variables. To assess differences in these variables by CVD risk category, Chi Squared tests were used for categorical variables and Kruskal-Wallis tests were used for numerical variables.

Unadjusted logistic regression models were fit to calculate weighted percentages of each outcome (MVP, other VHD, and any VHD) by CVD risk category, using sampling weights developed by the parent RURAL study. These weights are calculated such that weighted estimates are representative of the US population in the counties where the RURAL participants are recruited. Unadjusted weighted logistic regression models were also run to assess the associations between participant characteristics of interest (age, sex, race, BMI, cholesterol, blood pressure, diabetes, and eGFR) and each outcome.

Two primary weighted multivariable logistic regression models were then run for each outcome with CVD risk category as the primary covariate of interest: the first model was adjusted for race (White vs. non-White), BMI, and county, and the second model was additionally adjusted for age and sex. Age and sex are included in the PREVENT category calculation, therefore we ran two separate models – one that adjusted for age and sex and one that did not – because of concern for multicollinearity. Weighted left-censored Tobit regression models were used to evaluate the associations between log-transformed NTproBNP and hsTnT and MVP severity (“No MVP”, “Not Severe MVP”, and “Severe MVP”), adjusting for age, sex, race, BMI, and county. Missing values were imputed using multiple imputation with 20 datasets by chained equations using the mice package in R, and all model results were pooled across the 20 imputed datasets (**Supplemental Methods**). For all analyses, the overall level of significance was set to α=0.05. Data analyses were performed using R version 4.3.2; http://cran.r-project.org/.

## Results

We included 2,621 participants: 1,368 (52.2%) from Mississippi, 427 (16.3%) from Alabama, and 826 (31.5%) from Louisiana. The median age was 49 years (IQR 39.9-57.6), 68.7% were women, and 46% were Black (**Table 1**). Median BMI was 31.8 kg/m^2^ (IQR 27.2-38.0). Few (23.5%) had a pre-existing diagnosis of diabetes and median eGFR was 98.4 ml/min/1.73 m^2^ (IQR 84.0-109). Weighted imputed MVP prevalence was 1.7% and the weighted imputed prevalence of other VHD was 10.3% (**Figure 1**). Among those with MVP only, median age was 47.1 years (IQR 41.1-52.4), 59% were women, and median BMI was 26.3 kg/m^2^ (IQR 23.4-30.7). Among those with other VHD only, the median age was 55.6 years (IQR 45.4-60.4), 73.4% were women, and median BMI was 31.4 kg/m^2^ (IQR 27.4-38.0).

**Figure 1.**
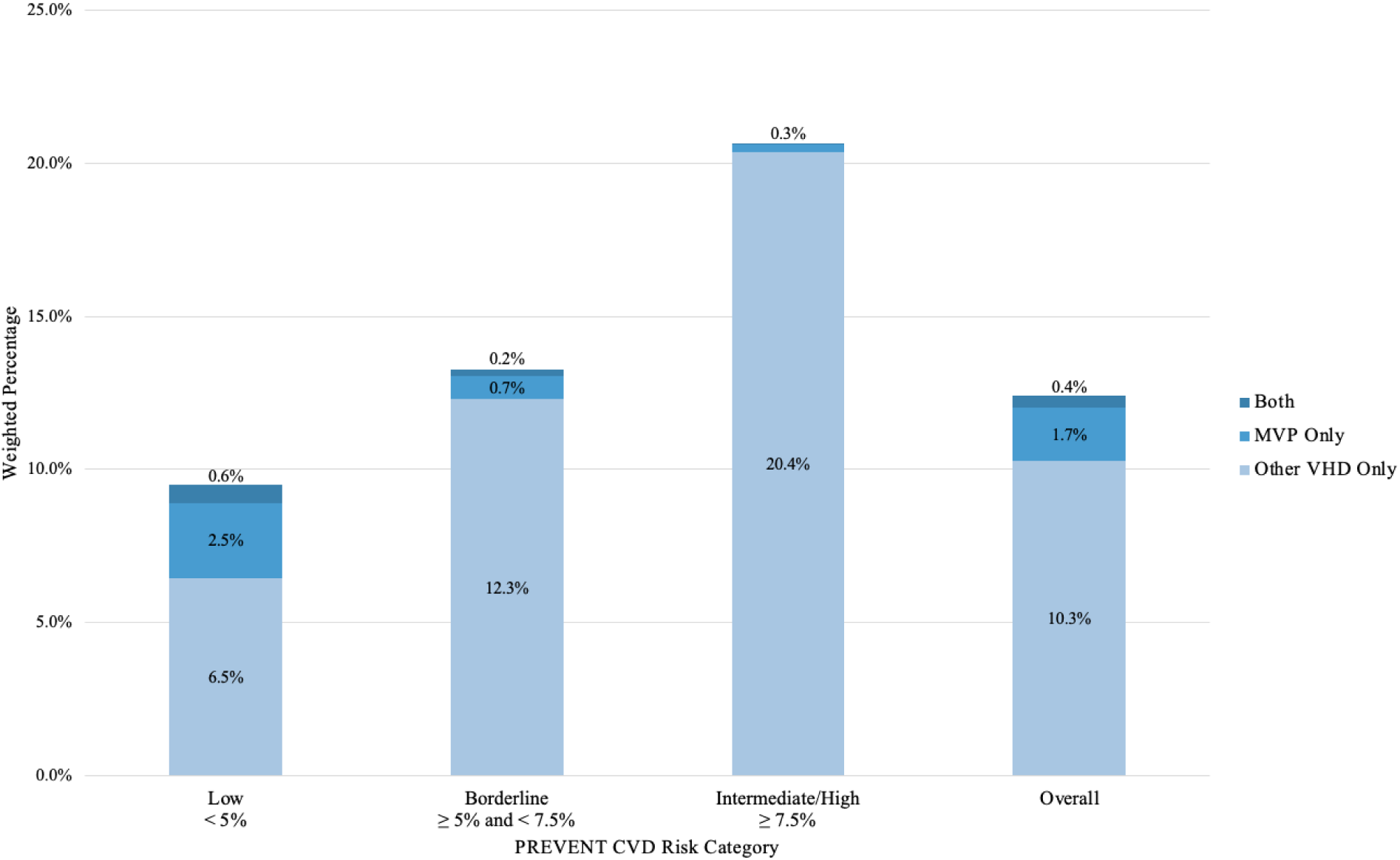
Weighted Imputed Percentages of MVP, other VHD, and both by CVD risk category Graphical depiction of the weighted imputed percentages of MVP, other VHD, and both categorized by PREVENT risk category and depicted overall. The percentage of both in the Intermediate/High risk group was 0%. CVD: cardiovascular disease; MVP: mitral valve prolapse; VHD: valvular heart disease.

**Table 1.** Baseline participant characteristics by estimated PREVENT CVD Risk Category.

| Characteristic | Overall<br>(N=2621) | Low<br>< 5%<br>(N=1455) | Borderline<br>≥ 5% and < 7.5%<br>(N=315) | Intermediate/High<br>≥ 7.5%<br>(N=561) | p-value |
| --- | --- | --- | --- | --- | --- |
| **Age (years), Median [Q1, Q3]** | 49.0 [39.9, 57.6] | 42.9 [35.8, 49.8] | 56.5 [50.6, 60.2] | 59.5 [54.8, 62.3] | <0.001 |
| **Female Sex, n (%)** | 1800 (68.7%) | 1049 (72.1%) | 214 (67.9%) | 327 (58.3%) | <0.001 |
| **BMI (kg/m<sup>2</sup>), Median [Q1, Q3]** | 31.8 [27.2, 38.0] | 30.7 [26.2, 37.8] | 32.5 [28.1, 38.0] | 33.3 [29.2, 38.5] | <0.001 |
| **Total Cholesterol (mg/dL), Median [Q1, Q3]** | 182 [158, 210] | 182 [160, 209] | 188 [154, 216] | 178 [154, 207] | 0.141 |
| **SBP (mmHg), Median [Q1, Q3]** | 124 [114, 136] | 119 [111, 129] | 130 [120, 142] | 137 [125, 151] | <0.001 |
| **DBP (mmHg), Median [Q1, Q3]** | 80.0 [73.0, 87.5] | 78.5 [72.0, 85.5] | 81.0 [73.3, 88.5] | 83.5 [75.0, 92.0] | <0.001 |
| **Diabetes Mellitus, n (%)** | 600 (23.5%) | 103 (7.1%) | 87 (27.6%) | 341 (60.8%) | <0.001 |
| **eGFR (ml/min/1.73 m<sup>2</sup>), Median [Q1, Q3]** | 98.4 [84.0, 109] | 102 [89.6, 112] | 92.6 [80.7, 104] | 89.1 [72.6, 100] | <0.001 |
| **CVD Risk Category, Median [Q1, Q3]** | 3.4 [1.2, 7.3] | 1.6 [0.7, 2.9] | 6.1 [5.6, 6.7] | 11.1 [9.0, 14.5] | <0.001 |
| **Race, n (%)** |  |  |  |  |  |
| American Indian<br>/Alaska Native | 2 (0.1%) | 0 (0%) | 1 (0.3%) | 0 (0.0%) |  |
| Asian | 16 (0.6%) | 5 (0.3%) | 4 (1.3%) | 2 (0.4%) |  |
| Native Hawaiian<br>or Other Pacific Islander | 1 (0.0%) | 1 (0.1%) | 0 (0.0%) | 0 (0.0%) |  |
| Black or African American | 1205 (46.0%) | 557 (38.3%) | 164 (52.1%) | 347 (61.9%) |  |
| White | 1363 (52.0%) | 867 (59.6%) | 141 (44.8%) | 210 (37.4%) |  |
| More than One Race | 4 (0.2%) | 2 (0.1%) | 1 (0.3%) | 0 (0.0%) |  |
| Unknown or<br>Not Reported | 30 (1.1%) | 23 (1.6%) | 4 (1.3%) | 2 (0.4%) |  |
| **MVP Present, n (%)** | 49 (1.9%) | 35 (2.5%) | 6 (2.0%) | 6 (1.1%) | 0.190 |
| **Other VHD Present, n (%)** | 293 (11.2%) | 126 (8.7%) | 45 (14.3%) | 88 (15.7%) | <0.001 |
| Summary statistics are unweighted. There were 290 individuals had missing CVD risk, 100 individuals with missing MVP status, 58 individuals with unknown total cholesterol, 1 individual missing average SBP, 1 individual missing average DBP, 72 individuals with unknown diagnosis of diabetes, and 60 individuals with unknown eGFR.<br>BMI: body mass index; CVD: cardiovascular disease; DBP: diastolic blood pressure; eGFR: estimated glomerular filtration rate; MVP: mitral valve prolapse; SBP: systolic blood pressure; VHD: valvular heart disease. |  |  |  |  |  |

Most individuals (1,455) were categorized as Low risk, 315 were Borderline risk, and 561 were Intermediate/High risk (**Table 1**). Median 10-year CVD risk was 1.6% (Low risk group), 6.1% (Borderline group), and 11.1% (Intermediate/High risk group). Intermediate/High risk individuals were older and had a higher BMI. The Intermediate/High risk group had fewer women (58.3% vs. 72.1% in Low-risk group) and more Black participants (61.9% vs. 38.3% in Low-risk group).

Among 2,621 participants, 49 had MVP, 293 had other VHD, 10 had both, and 100 (4%) were missing MVP status. **Supplemental Tables 1 and 2** report the echocardiographic findings of those with other VHD. Most other VHD were mitral annular calcification alone (59%); isolated aortic abnormalities were the second-most common (13%).

MVP and other VHD prevalence differed by CVD risk. MVP weighted imputed prevalence was 2.5% in the Low risk group vs. 0.3% in Intermediate/High risk group. Other VHD was prevalent in 6.5% in the Low risk group vs. 20.4% in Intermediate/High risk group (**Figure 1**). Age was inversely associated with odds of MVP (OR 0.56, 95% CI 0.47-0.67) and positively associated with the odds of other VHD and any VHD (**Table 2**). White race was associated with higher odds of MVP (OR 3.07, 95% CI 1.75-5.41), other VHD (OR 1.38, 95% CI 1.25-1.51), and any VHD (OR 1.55, 95% CI 1.36-1.76). Female sex was associated with lower odds of MVP (OR 0.56, 95% CI 0.36-0.86), slightly higher odds of other VHD (OR 1.12, 95% CI 1.04-1.21), and lacked a significant association with any VHD. Higher BMI was associated with lower odds of MVP (OR 0.48, 95% CI 0.36-0.65).

**Table 2.** Associations between Participant Characteristics and MVP/VHD.

| Characteristic | MVP |  | Other VHD |  | Any VHD |  |
| --- | --- | --- | --- | --- | --- | --- |
|  | OR (95% CI) | p-value | OR (95% CI) | p-value | OR (95% CI) | p-value |
| **Age (per 11.78 years)** | 0.56 (0.47, 0.67) | <0.001 | 1.92 (1.84, 2.00) | <0.001 | 1.62 (1.51, 1.74) | <0.001 |
| **White** |  |  |  |  |  |  |
| **No** | Ref |  | Ref |  | Ref |  |
| **Yes** | 3.07 (1.75, 5.41) | <0.001 | 1.38 (1.25, 1.51) | <0.001 | 1.55 (1.36, 1.76) | <0.001 |
| **Sex** |  |  |  |  |  |  |
| **Male** | Ref |  | Ref |  | Ref |  |
| **Female** | 0.56 (0.36, 0.86) | 0.010 | 1.12 (1.04, 1.21) | 0.004 | 1.03 (0.92, 1.15) | 0.66 |
| **BMI (per 8.3 kg/m<sup>2</sup>)** | 0.48 (0.36, 0.65) | <0.001 | 1.07 (1.03, 1.12) | <0.001 | 1.01 (0.96, 1.07) | 0.735 |
| **Total Cholesterol (per 41.94 mg/dL)** | 1.24 (1.15, 1.34) | <0.001 | 1.13 (1.09, 1.17) | <0.001 | 1.13 (1.09, 1.17) | <0.001 |
| **Average SBP (per 17.58 mmHg)** | 0.83 (0.73, 0.93) | 0.003 | 1.28 (1.24, 1.33) | <0.001 | 1.23 (1.18, 1.28) | <0.001 |
| **Average DBP (per 11.42 mmHg)** | 0.95 (0.77, 1.18) | 0.623 | 0.92 (0.87, 0.97) | 0.003 | 0.93 (0.88, 0.98) | 0.008 |
| **Diabetes Mellitus** |  |  |  |  |  |  |
| **No** | Ref |  | Ref |  | Ref |  |
| **Yes** | 0.44 (0.28, 0.70) | <0.001 | 2.24 (2.03, 2.47) | <0.001 | 1.99 (1.81, 2.19) | <0.001 |
| **eGFR (per 18.95 mL/min/1.73m<sup>2</sup>)** | 1.71 (1.55, 1.89) | <0.001 | 0.77 (0.73, 0.80) | <0.001 | 0.83 (0.79, 0.87) | <0.001 |

\*ORs for continuous variables are presented for a change of one estimated standard deviation in the counties of interest.
BMI: body mass index; CI: confidence interval; DBP: diastolic blood pressure; eGFR: estimated glomerular filtration rate; MVP: mitral valve prolapse; OR: odds ratio; Ref: referent; SBP: systolic blood pressure; VHD: valvular heart disease.

In both our our minimally adjusted (Model 1) and multivariable-adjusted (Model 2) models, MVP odds were lower in the Borderline risk group (OR 0.34, 95% CI 0.19-0.58 in Model 1 and OR 0.51, 95% CI 0.34-0.78 in Model 2) and Intermediate/High risk group (OR 0.13, 95% CI 0.09-0.19 and OR 0.22, 95% CI 0.13-0.38) than in the Low risk group (**Figure 2** top two rows, **Table 3**). The odds of having other VHD were higher in the Intermediate/High risk group in both models (OR 3.77, 95% CI 3.36-4.23 and OR 2.06, 95% CI 1.73-2.44) (**Figure 2** middle two rows).

**Figure 2.**
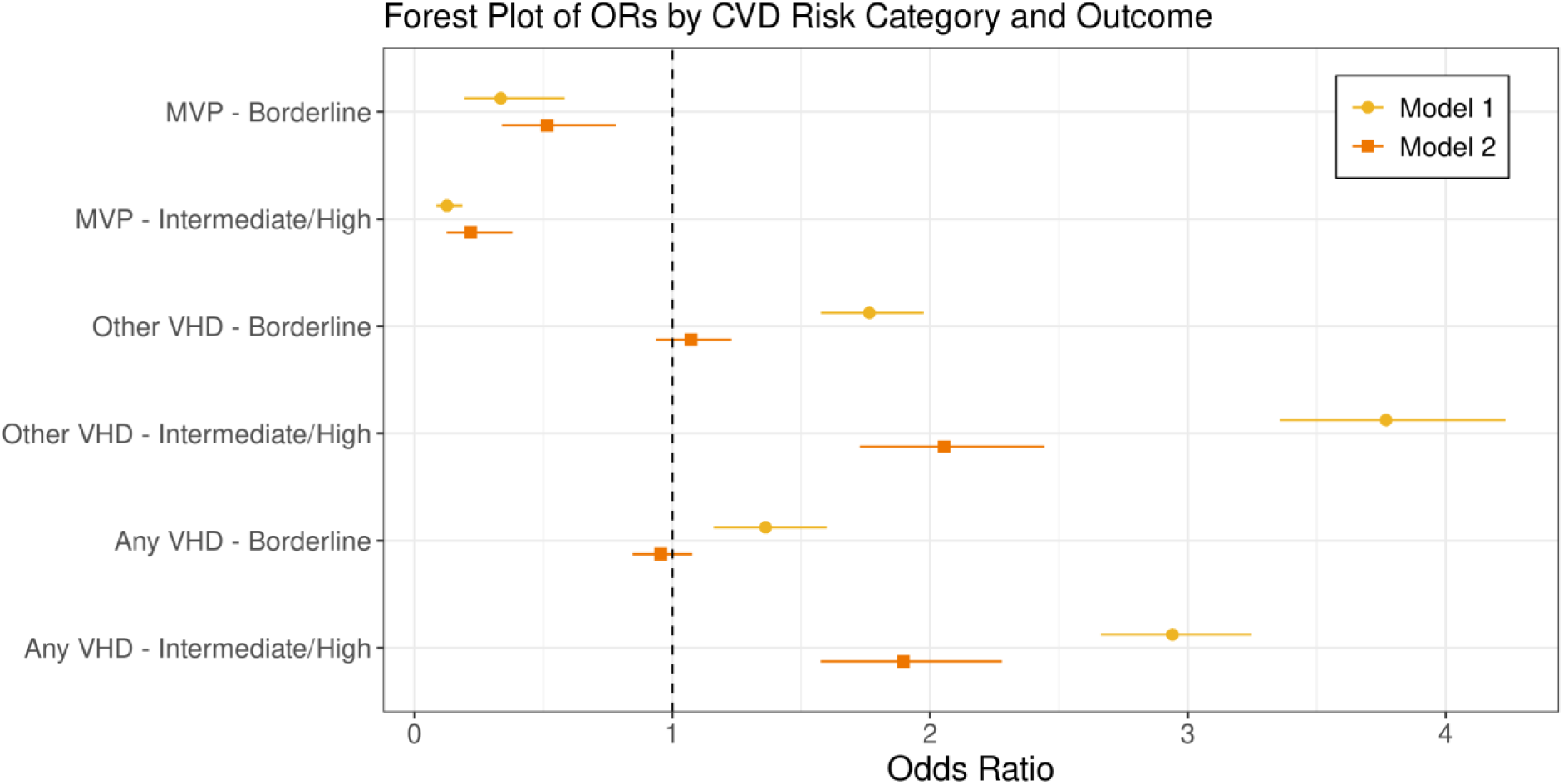
Forest Plot of Odds Ratios by CVD Risk Category and Outcome Depiction of odds ratios for MVP, other VHD, and any VHD by CVD risk category. Includes odds ratios from both our minimally adjusted (Model 1) and multivariable-adjusted (Model 2) models. CVD: cardiovascular disease; MVP: mitral valve prolapse; VHD: valvular heart disease

**Table 3.** Association between CVD Risk Category, MVP and VHD.

| Outcome & CVD Risk Category | Model 1 |  | Model 2 |  |
| --- | --- | --- | --- | --- |
|  | OR (95% CI) | p-value | OR (95% CI) | p-value |
| **MVP** |  | <0.001 |  | <0.001 |
| **Low** | Ref |  | Ref |  |
| **Borderline** | 0.34 (0.19, 0.58) | <0.001 | 0.51 (0.34, 0.78) | 0.002 |
| **Intermediate/High** | 0.13 (0.09, 0.19) | <0.001 | 0.22 (0.13, 0.38) | <0.001 |
| **Other VHD** |  | <0.001 |  | <0.001 |
| **Low** | Ref |  | Ref |  |
| **Borderline** | 1.77 (1.58, 1.98) | <0.001 | 1.07 (0.94, 1.23) | 0.31 |
| **Intermediate/High** | 3.77 (3.36, 4.23) | <0.001 | 2.06 (1.73, 2.44) | <0.001 |
| **Any VHD** |  | <0.001 |  | <0.001 |
| **Low** | Ref |  | Ref |  |
| **Borderline** | 1.36 (1.16, 1.60) | <0.001 | 0.95 (0.85, 1.08) | 0.45 |
| **Intermediate/High** | 2.94 (2.67, 3.25) | <0.001 | 1.90 (1.58, 2.28) | <0.001 |
| Model 1 is adjusted for Race (White vs. Non White), BMI, and County. Model 2 is adjusted for Age, Sex, Race (White vs. Non White), BMI, and County. A third model (not shown) was developed for this analysis that controlled for county, age, sex and BMI that yielded substantively similar results as Model 2.<br>BMI: body mass index; CI: confidence interval; CVD: cardiovascular disease; MVP: mitral valve prolapse; OR: odds ratio; Ref: referent; VHD: valvular heart disease. |  |  |  |  |

Compared to the Low risk group, odds for any VHD were higher in both Borderline and Intermediate/High risk groups in the minimally-adjusted models, whereas only the odds for the Intermediate/High risk group remained significant after multivariable adjustment (OR 1.90, 95% CI 1.58-2.28) (**Figure 2** bottom two rows, **Table 3**). A third model was developed for this analysis that controlled for county, age, sex, and BMI that yielded substantively similar results as the second model.

Among a subset of 1,000 participants with biomarker and adequate echocardiogram data, hsTnT was undetectable (i.e., below assay minimal detection limits, MDL) in 677 participants (MDL < 6 ng/L) and NTproBNP was undetectable in 564 participants (MDL < 36 pg/mL). None of the participants with severe MVP had detectable hsTnT values. Both biomarker levels were lower among those with non-severe MVP compared to those without MVP. NTproBNP levels were 2.05 times greater (95% CI 1.49-2.83) in participants with severe MVP vs. without MVP (**Figure 3**, **Table 4**).

**Figure 3.**
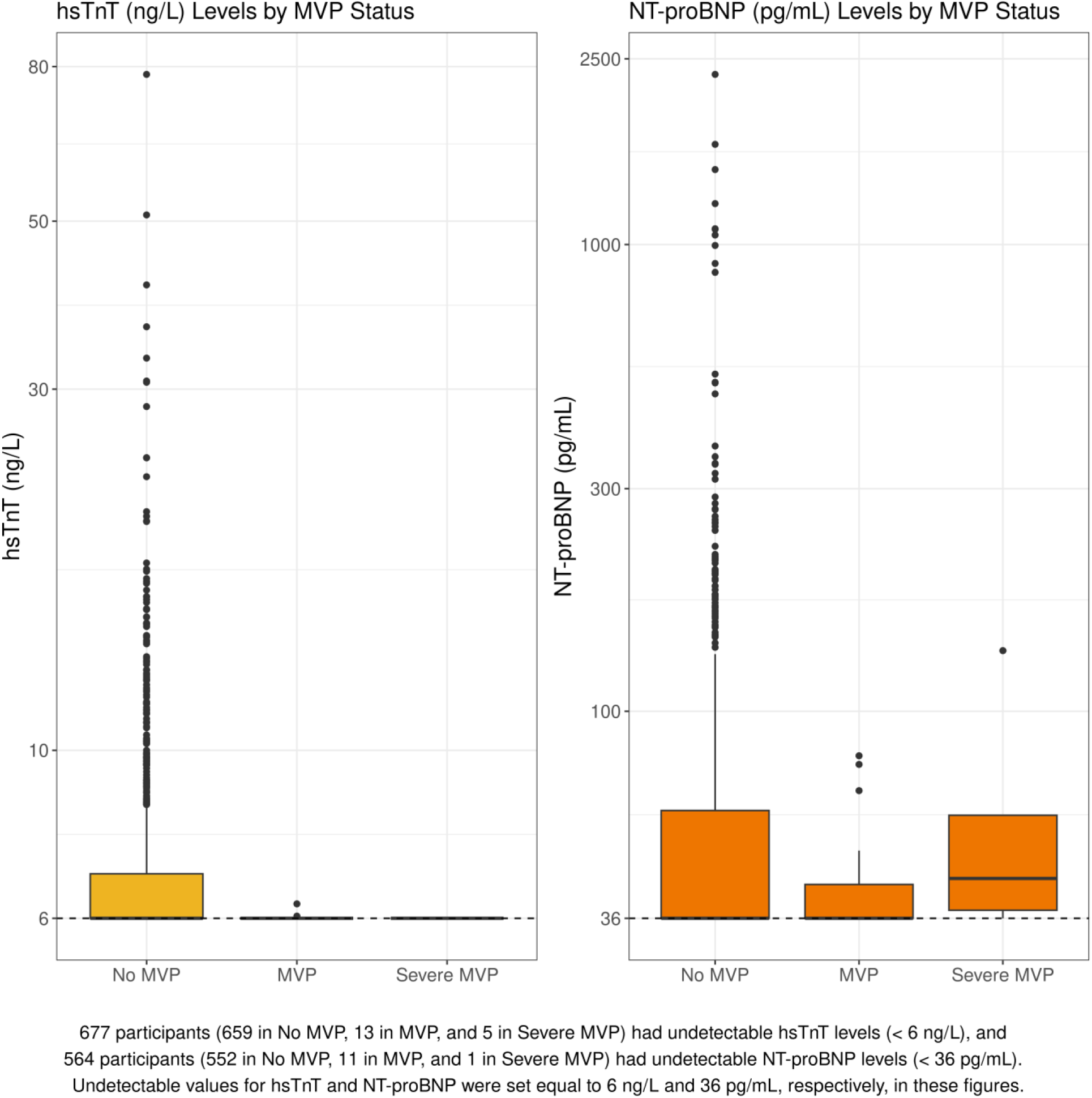
Biomarker Levels by MVP Status Boxplot demonstrating biomarker levels according to MVP status. hsTnT: high-sensitivity cardiac troponin T; MVP: mitral valve prolapse; NTproBNP: N-terminal pro b-type natriuretic peptide

**Table 4.**
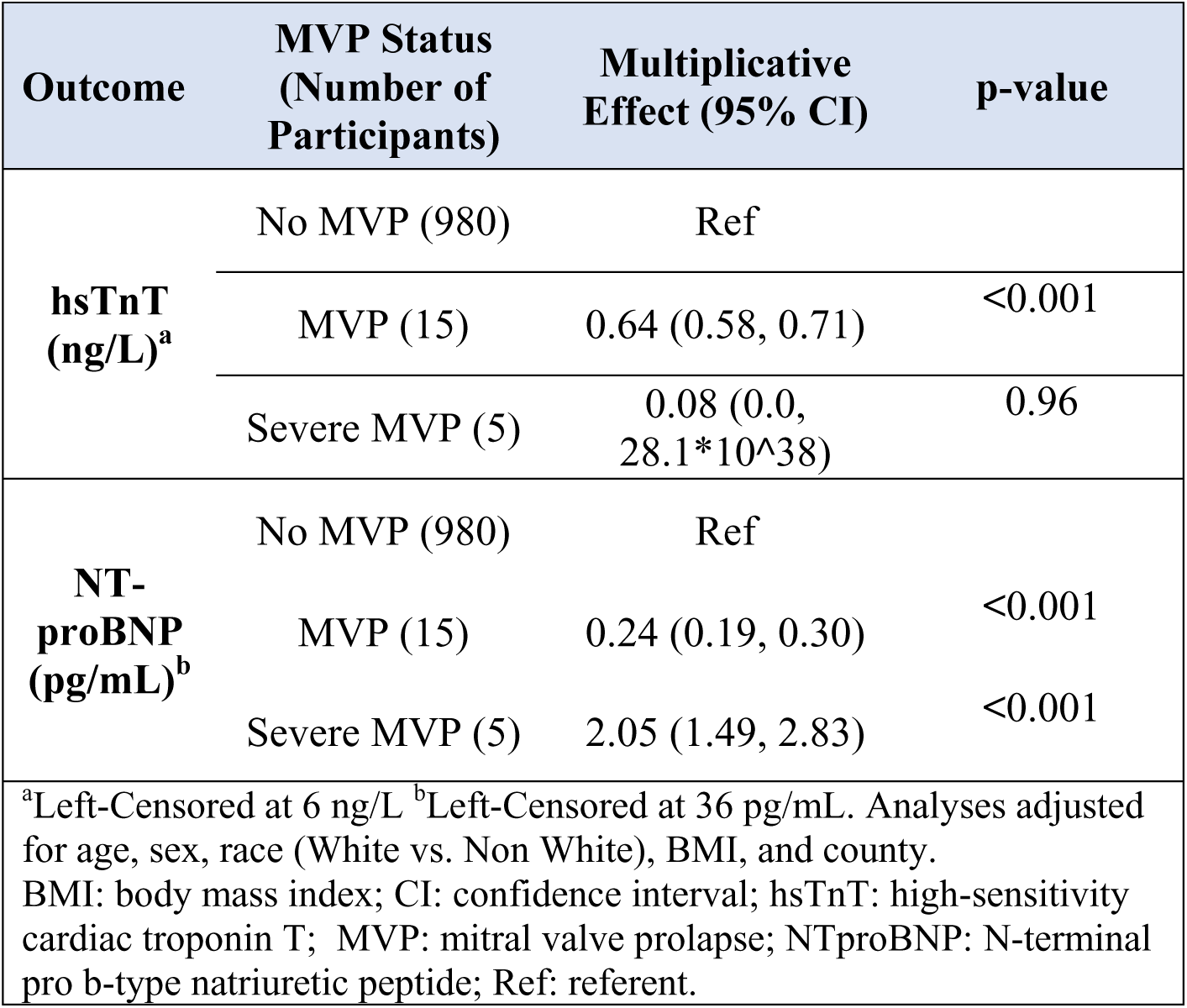
Associations between MVP Severity and Biomarker Values.

| **Outcome** | **MVP Status**<br>**(Number of**<br>**Participants)** | **Multiplicative**<br>**Effect (95% CI)** | **p-value** |
| --- | --- | --- | --- |
| **hsTnT**<br>**(ng/L)<sup>a</sup>** | No MVP (980) | Ref |  |
|  | MVP (15) | 0.64 (0.58, 0.71) | <0.001 |
|  | Severe MVP (5) | 0.08 (0.0,<br>28.1*10<sup>38</sup>) | 0.96 |
| **NT-**<br>**proBNP**<br>**(pg/mL)<sup>b</sup>** | No MVP (980) | Ref |  |
|  | MVP (15) | 0.24 (0.19, 0.30) | <0.001 |
|  | Severe MVP (5) | 2.05 (1.49, 2.83) | <0.001 |
| <sup>a</sup>Left-Censored at 6 ng/L <sup>b</sup>Left-Censored at 36 pg/mL. Analyses adjusted for age, sex, race (White vs. Non White), BMI, and county.<br>BMI: body mass index; CI: confidence interval; hsTnT: high-sensitivity cardiac troponin T; MVP: mitral valve prolapse; NTproBNP: N-terminal pro b-type natriuretic peptide; Ref: referent. |  |  |  |

## Discussion

We performed echocardiography and collected detailed medical histories to evaluate MVP/VHD prevalence, risk associations, and biomarker relationships in the rural South. Our principal findings are (1) MVP prevalence was consistent with prior population-based studies, (2) higher MVP/VHD odds were associated with White race, while lower MVP odds were linked to increasing age, female sex, and BMI, (3) VHD odds rose with CVD risk, while MVP odds declined, and (4) NTproBNP levels were higher in severe MVP compared to those without MVP. These findings add new insights from under-represented rural areas of the US on VHD patterms.

In our sample from five rural counties, the prevalence of MVP was comparable to that in historical large cohort studies.^9,11,12^ The RURAL cohort study population includes a broader representation of a diverse group of Americans across important socio-demographic groups for whom little data are available. The median age of our cohort was 49, younger than participants in the Strong Heart Study whose ages ranged from 45-74 years^11^ and older than participants in the CARDIA study with ages between 23-35 years.^25^ The Framingham Heart Study (FHS) found that age distributions of those with MVP were similar to those without MVP; participant ages were between 26-84 years old.^9^ Despite these age differences, the prevalence of MVP appears consistently ∼2%.

Our sample includes more Black individuals compared to previous large cohorts. An analysis from the Coronary Artery Risk Development Study (CARDIA), one of the only studies to specifically address potential differences in the burden of MVP among Black Americans, reported no significant difference in prevalence compared to White Americans.^25^ In our analysis, White individuals had higher odds of MVP, other VHD, and any VHD, though the small sample size of MVP restricts generalization of this finding. MVP is characterized by familial clustering and high heritability.^26^ Prevalence estimates for MVP have been fairly consistent across racially and ethnically diverse studies, including the Framingham Offspring Heart Study predominantly involving White participants, the Strong Heart Study involving American Indian participants, and a multiethnic population-based study consisting of South Asian, European, and Chinese participants in Canada.^11,12^ Thus, the variation in odds of MVP by race in our sample is more likely to reflect imprecision of point estimates due to a modest sample size as opposed to a true difference based on race, which is a social construct.

Previous results have been mixed on the veracity of a female sex predilection for MVP. The prevailing paradigm suggests that the prevalence of MVP is greater in women than men^26,27^ but not all studies support a sex difference.^9,11^ In our population, women had lower odds of MVP relative to men which stands in contrast to most of the prior literature. It may be that women in the rural US South are less likely than men in the region to have MVP, an observation that could be related to ancestry and heritable traits in the region that have not yet been identified. Human and animal studies of MVP have shown some forms of MVP to be primarily autosomal dominantly inherited with incomplete and age-dependent penetrance or an X chromosome inherited disease.^28^ We are not aware of studies from the rural US South that have specifically explored these potential pathways and we suggest larger studies to further evaluate and replicate the interactions between MVP, region, and sex.

In our analysis, higher BMI was associated with lower odds of MVP. The relationship between BMI and MVP/VHD has been previously evaluated with mixed findings, though inverse relationships have been noted using Mendelian randomization studies on individuals of European ancestry.^29^ The observed relationship between lower BMI and higher prevalence of MVP may be better explained by easier echocardiographic detection of abnormalities in individuals with lower BMI as opposed to a mediating effect of body fat storage or composition predispositions. Younger individuals may be leaner, allowing easier detection of MVP by echo. Together the observations about BMI and MVP suggest the presence of distinct pathology in MVP compared to other VHD and further study into these associations is merited.

The combined prevalence of other VHD in our sample was just above 10%. The Atherosclerosis Risk in Communities (ARIC) study found that 38.6% of individuals were “at risk” for further valve dysfunction and 16.5% had progressive VHD.^30^ The mean age of ARIC participants was 76 years^30^, notably older than our study population. Regurgitant lesions have comprised most of the VHD in contemporary population-based epidemiological studies, and the incidence and severity of valvular regurgitation is postulated to increase with age.^10,31–34^ In the current study, increasing age was associated with approximately double the odds of VHD. Our study only quantified those with moderate or greater disease as having VHD which may explain the differences observed between our study and other large cohorts. We observed a small increase in odds for other VHD associated with increasing BMI yet did not observe increased odds for any VHD, an inconsistent result, which reflects the prior literature.^10,35^

The extent to which the AHA’s PREVENT equations’ 10-year CVD risk estimates associate with VHD has been incompletely understood. After multivariable adjustment for relevant covariates including BMI, we found different associations between MVP, VHD, and estimated CVD risk; higher predicted risk was associated with greater odds of VHD and lower odds of MVP. Most VHD are atherosclerotic or degenerative lesions that share risk factors with those included in the PREVENT risk estimation. While we adjusted for factors including BMI, it remains possible that differences in body composition may account for the observed association. BMI does not take into account anthropometric differences such as distributions of fat, skeletal, muscle, or chest dimensions that could both related to estimated CVD risk and ability to identify MVP by echo. We also note that the observed association between higher risk classification and a lower odds for MVP may result from the modest sample size of those with MVP, making this association uncertain. This relationship persisted in both of our adjusted models. This prompts the need for further research into addressing common social or environmental exposures in rural communities and VHD risk as we intend to pursue in the full cohort. There is also a need for further research into appropriate risk calculations for MVP and VHD – in patients who might lack traditional CVD risk factors or for whom these risk factors are not predictive of disease onset and prognosis.

We sought to elucidate the association between MVP severity and cardiac biomarkers in a subsample of our cohort. MVP was associated with a lower hsTnT, an indicator of myocardial injury, and we did not observe a relationship between severe MVP and hsTnT. We would anticipate that hsTnT increases in conditions associated with hypoxia-induced injury^36^, and would not necessarily expect the same result in those with MVP, which is a structural abnormality related to proteoglycan accumulation as opposed to a predominantly ischemic process.^37^ We observed that circulating NTproBNP concentrations were significantly higher in participants with severe MVP compared with participants without MVP, though NTproBNP concentrations were significantly lower in those with non-severe MVP compared to those without MVP. Circulating NTproBNP concentrations are positively associated with the incidence of heart failure and CVD death.^38^ In a community-based sample from the FHS, higher levels of NTproBNP carried a 77% increase in the risk of heart failure and a 27% increase in the risk of death.^36^ Similar findings have been reported in the Multi-Ethnic Study of Atherosclerosis (MESA) as well as in international cohorts.^20,21^ As severe MVP may progress to mitral regurgitation, higher circulating NTproBNP concentrations may reflect worsening left atrial or ventricular stretch. Our finding of significantly higher NTproBNP associated with severe MVP may have clinical implications for risk stratifying MVP, especially in regions with limited echocardiographic imaging or in patients in whom echocardiography is technically challenging. Indeed, high-risk MVP phenotypes were the impetus for the national investment (i.e., CAROL Act) in disentangling features of MVP that indicate progressive disease.

The strengths of our investigation include utilizing novel AI-guided echocardiography, a method that is feasible and reproducible in communities facing barriers to access to cardiac imaging. We also relied on core lab-defined cases of MVP and associated echocardiographic findings. By nature of the design of the RURAL study, we also achieved representation across individual CVD risk categories for our analysis. Several limitations of our investigation are also worth noting. We had a small number of individuals with MVP in our sample, which reduced the precision of our estimates of risk and associations with participant characteristics. Furthermore, we only included individuals who had sufficient quality echo images for identifying MVP and VHD, possibly missing cases of MVP that were not able to be identified by transthoracic echo. By nature of our AI-echo acquisition protocol, we were not able to include spectral Doppler data in RURAL’s Exam 1. As a result, we are limited in assessing diastolic function and hemodynamic assessments. By the nature of the study design of this supplemental study, we were limited to the subset of individuals for whom we could perform biomarker testing, which limits the generalizability of our results. Notably, the instability of the estimates may be due to low counts of severe MVP and large degree of censoring due to undetectable biomarker levels.

## Conclusions

Using a novel MEU approach in RURAL, our study demonstrates that MVP and other VHD contribute to the burden of CVD in rural counties in the southern US. The association between the higher CVD risk category and the odds of any VHD requires further evaluation. We identify that NTproBNP levels may identify individuals with MVP with more severe features. Future studies in this or other cohorts are needed to confirm whether these associations are associated with adverse cardiac outcomes. Such data are a critical next step to mitigate the disproportionate CVD burden in rural America.

## Clinical Perspectives

The RURAL study aims to address barriers to care in rural areas. Using AI-guided echocardiography in MEUs proves a novel way to reach rural communities previously not involved in major CVD research efforts. Our study found that a higher CVD risk may be associated with certain forms of VHD and that severe MVP is associated with higher levels of NTproBNP. These findings support efforts to enhance screening and treatment of valvular heart disease in rural areas.

## Abbreviation List

AI: Artificial intelligence
MVP: Mitral valve prolapse
VHD: Valvular heart disease
CVD: Cardiovascular disease
MEUs: Mobile examination units
RURAL: Echo Risk Underlying Rural Areas Longitudinal Echocardiographic Ancillary Study
PREVENT: Predicting Risk of CVD EVENTs
hsTnT: High-sensitivity cardiac troponin T
NTproBNP: N-terminal pro-B-type natriuretic peptide

## Data Availability

All data produced in the present study are available upon reasonable request to the authors

## Acknowledgments

The investigators acknowledge the contribution of the RURAL Cohort Study participants, who continue to make this research possible and Caption Health (GE) for in-kind support for AI-echos.

---

Central Illustration: Bringing echo to rural communities facilitates risk prediction evaluation and cardiac biomarker/disease severity analysis (A) Map indicating states included in our analysis. (B) A higher PREVENT risk category increases odds for having any VHD. (C) NTproBNP levels were higher in those with severe MVP compared to those without MVP. CVD: cardiovascular disease; MVP: mitral valve prolapse; NTproBNP: N-terminal pro b-type natriuretic peptide.

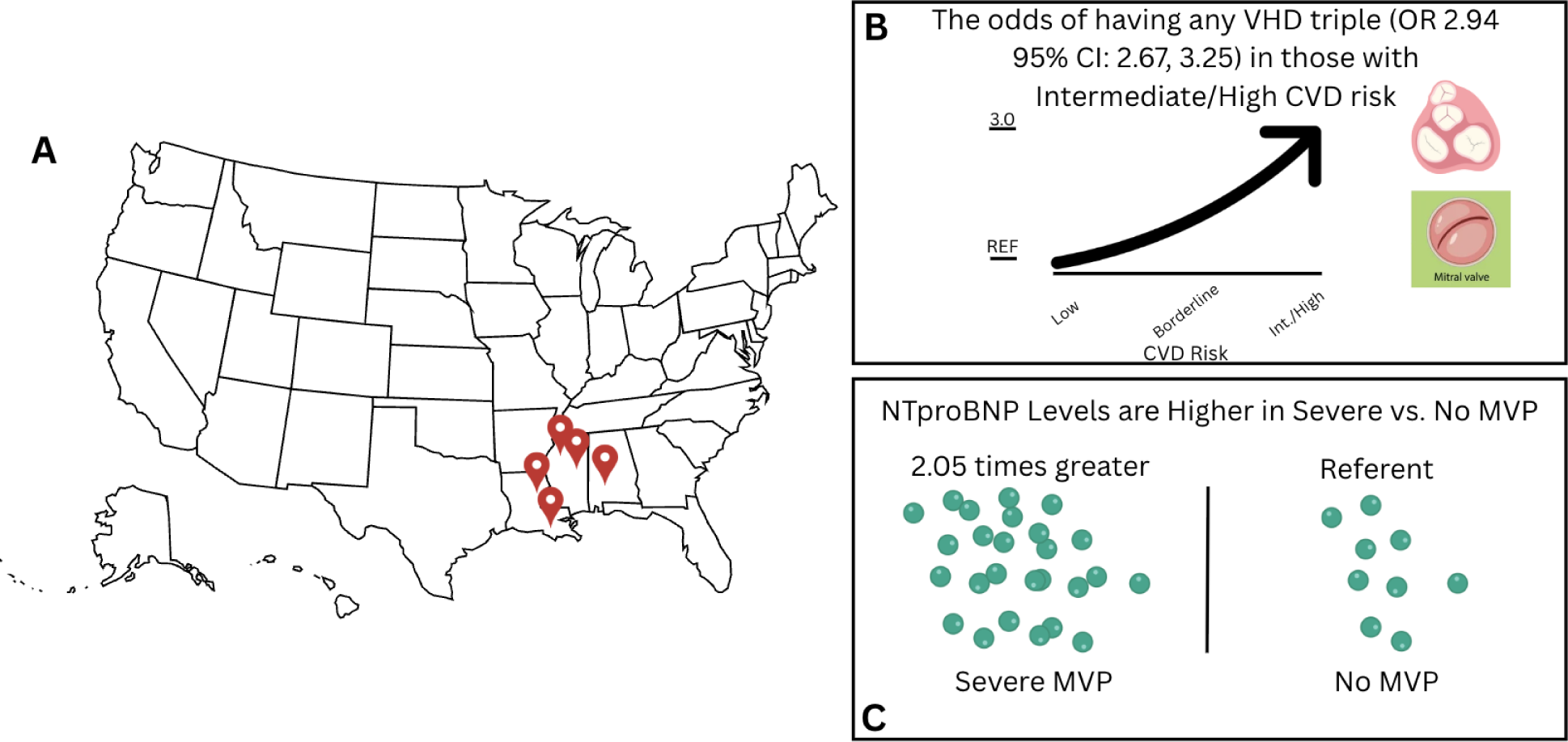

