## Supplementary Material for "Valvular Heart Disease Associations with Cardiac Biomarkers Using AI-guided Echocardiography: the RURAL Cohort Study"

### Supplemental Methods

*Image Acquisition Using AI-guided Echo*

The software relies exclusively on ultrasound images and does not require additional trackers or sensors. Six RURAL technicians (nonsonographers) in the MEU use AI-powered, real-time feedback and guidance during image acquisition to ensure optimal transducer positioning, probe angle, and equipment settings adjustments, resulting in clearer and diagnostically valuable cardiac images. The 2-dimensional echo images were obtained in the parasternal long- and short-axis views, as well as apical 4-, 2-, and 3-chamber views. Color Doppler was acquired for evaluation of valvular regurgitation. All images were acquired using Terason uSmart 3200t Plus Ultrasound System with Caption software (version 0.1.0.1654184491_ECG) at 60 to 90 frames/sec and digitally archived.

*Echocardiographic Measurements for Left Ventricular Ejection Fraction and Global Longitudinal Strain*

Left ventricular ejection fraction (LVEF) was measured using the biplane Simpson volumetric method combining apical 4- and 2-chamber views. LVEF was also determined by visual estimation (in 5-point increments). When the definition of the LV endocardial border was not adequate for biplane tracing, visual LVEF substituted to provide a single combined LVEF determination in all patients. For the present analysis, LV dysfunction was defined as an LVEF < 50%. Global longitudinal strain was performed on apical views in the DCRI ICL, and chamber dimensions were measured in the parasternal long and apical views.

*Cardiac Biomarker Assays*

HsTnT and NTproBNP were measured in serum collected at the baseline participant visit. All analyses were performed at the RURAL Biorepository and Clinical Biochemistry Core Lab (University of Vermont, Burlington, VT, USA). HsTnT was measured using the Elecsys Troponin T Gen 5 STAT assay system (Roche Diagnostics, Indianapolis, IN, USA).[1] NTproBNP was measured using Abcam's BNP Human ELISA Kit (Abcam, Cambridge, United Kingdom).[2]

*Description of Imputation Model Variables*

Variables in the imputation model included MVP, an indicator for echocardiographic abnormalities, all 10 variables that contribute to other VHD, other VHD itself, all variables included in the PREVENT risk score, demographic characteristics (age, race, sex, BMI, and county), diastolic blood pressure, both biomarkers with undetectable values set to the minimal detection limits, and left atrial dimension, volume and systolic area. Missing values were imputed using multivariate imputation by chained equations using the mice package in R. The default mice imputation methods were used (continuous variables were imputed using predictive mean matching and categorical variables were imputed using logistic regression), and 20 imputed datasets were created. Models were fit within each imputed dataset, and pooled model estimates (coefficients, standard errors, and p-values) were obtained using Rubin's rules.[3]

**Supplemental Table 1. Distribution of other VHD Echocardiographic Characteristics by CVD Risk Category***

| Characteristic | Overall (N=2621) | Low (N=1455) | Borderline (N=315) | Intermediate/ High (N=561) |
|---|---|---|---|---|
| **MR** | | | | |
| **Moderate to Severe MR** | 6 (0.3%) | 3 (0.3%) | 0 (0.0%) | 3 (0.9%) |
| **None** | 885 (50.0%) | 526 (51.9%) | 103 (48.8%) | 173 (49.7%) |
| **Present Unable to quantitate** | 38 (2.1%) | 18 (1.8%) | 8 (3.8%) | 6 (1.7%) |
| **Trivial to Mild MR** | 841 (47.5%) | 466 (46.0%) | 100 (47.4%) | 166 (47.7%) |
| **Missing** | 851 | 442 | 104 | 213 |
| **TR** | | | | |
| **Moderate to Severe TR** | 1 (0.3%) | 0 (0.0%) | 1 (2.3%) | 0 (0.0%) |
| **None** | 166 (41.7%) | 97 (40.9%) | 15 (34.1%) | 33 (44.0%) |
| **Present Unable to quantitate** | 63 (15.8%) | 36 (15.2%) | 8 (18.2%) | 15 (20.0%) |
| **Trivial to Mild TR** | 168 (42.2%) | 104 (43.9%) | 20 (45.5%) | 27 (36.0%) |
| **Missing** | 2223 | 1218 | 271 | 486 |
| **AR** | | | | |
| **Moderate to Severe AR** | 2 (0.2%) | 1 (0.1%) | 0 (0.0%) | 0 (0.0%) |
| **None** | 1194 (93.6%) | 692 (94.9%) | 147 (95.5%) | 220 (89.8%) |
| **Present Unable to quantitate** | 6 (0.5%) | 0 (0%) | 1 (0.6%) | 5 (2.0%) |

| Characteristic | Overall (N=2621) | Low (N=1455) | Borderline (N=315) | Intermediate/ High (N=561) |
|---|---|---|---|---|
| **Trivial to Mild AR** | 74 (5.8%) | 36 (4.9%) | 6 (3.9%) | 20 (8.2%) |
| **Missing** | 1345 | 726 | 161 | 316 |
| **Congenital Abnormalities** | | | | |
| **Absent** | 819 (99.9%) | 462 (99.8%) | 100 (100.0%)) | 174 (100.0%) |
| **Present** | 1 (0.1%) | 1 (0.2%) | 0 (0.0%) | 0 (0.0%) |
| **Missing** | 1801 | 992 | 215 | 387 |
| **AV** | | | | |
| **Mild to Moderate Structural Abnormality** | 70 (3.2%) | 20 (1.6%) | 12 (4.7%) | 27 (6.0%) |
| **Normal** | 2089 (96.4%) | 1193 (98.1%) | 242 (95.3%) | 420 (93.3%) |
| **Other** | 5 (0.2%) | 1 (0.1%) | 0 (0.0%) | 3 (0.7%) |
| **Severe Structural Abnormality** | 2 (0.1%) | 2 (0.2%) | 0 (0.0%) | 0 (0.0%) |
| **Missing** | 455 | 239 | 61 | 111 |
| **TV** | | | | |
| **Mild to Moderate Structural Abnormality** | 2 (0.1%) | 1 (0.1%) | 0 (0%) | 0 (0%) |
| **Normal** | 1780 (99.9%) | 1022 (99.9%) | 211 (100%) | 347 (100%) |
| **Missing** | 839 | 432 | 104 | 214 |
| **MV** | | | | |
| **Mild to Moderate** | 46 (1.8%) | 18 (1.2%) | 9 (2.9%) | 13 (2.4%) |

| Characteristic | Overall (N=2621) | Low (N=1455) | Borderline (N=315) | Intermediate/ High (N=561) |
|---|---|---|---|---|
| **Structural Abnormality** | | | | |
| **Normal** | 2545 (98.1%) | 1422 (98.7%) | 304 (97.1%) | 537 (97.5%) |
| **Other** | 1 (0.0%) | 1 (0.1%) | 0 (0.0%) | 0 (0.0%) |
| **Severe Structural Abnormality** | 1 (0.0%) | 0 (0.0%) | 0 (0.0%) | 1 (0.2%) |
| **Missing** | 28 | 14 | 2 | 10 |
| **AV Structure** | | | | |
| **Bicuspid** | 1 (0.0%) | 0 (0.0%) | 0 (0.0%) | 0 (0.0%) |
| **Normal** | 2152 (82.1%) | 1211 (83.2%) | 254 (80.6%) | 444 (79.1%) |
| **Other Abnormal** | 9 (0.3%) | 4 (0.3%) | 0 (0.0%) | 4 (0.7%) |
| **Unable to be assessed** | 459 (17.5%) | 240 (16.5%) | 61 (19.4%) | 113 (20.1%) |
| **MAC** | | | | |
| **Absent** | 2381 (91.9%) | 1349 (93.6%) | 279 (89.4%) | 489 (88.7%) |
| **Present** | 211 (8.1%) | 92 (6.4%) | 33 (10.6%) | 62 (11.3%) |
| **Missing** | 29 | 14 | 3 | 10 |
| **Heart Valves** | | | | |
| **Normal** | 1764 (67.3%) | 1011 (69.5%) | 198 (62.9%) | 357 (63.6%) |
| **Thickened** | 16 (0.6%) | 5 (0.3%) | 1 (0.3%) | 7 (1.2%) |
| **Unable to be assessed** | 841 (32.1%) | 439 (30.2%) | 116 (36.8%) | 197 (35.1%) |

*Missing observations are not included in the denominator of the reported percentages.

AR: aortic regurgitation; AV: aortic valve; MAC: mitral annular calcification; MR: mitral regurgitation; MV: mitral valve; TR: tricuspid regurgitation; TV: tricuspid valve

**Supplemental Table 2. Combinations of echocardiographic findings among the 293 participants with Other VHD.**

| Condition | Count (%) |
|---|---|
| **Mitral Annular Calcification** | 173 (59.0%) |
| **AV Structural Abnormality** | 38 (13.0%) |
| **MV Structural Abnormality** | 21 (7.2%) |
| **AV Structural Abnormality & Mitral Annular Calcification** | 15 (5.1%) |
| **MV Structural Abnormality & Mitral Annular Calcification** | 10 (3.4%) |
| **AV Structural Abnormality & MV Structural Abnormality** | 6 (2.1%) |
| **AV Structural Abnormality & Abnormal Structure of AV & Thickened Heart Valves** | 4 (1.4%) |
| **MV Structural Abnormality & Mitral Annular Calcification & Thickened Heart Valves** | 3 (1.0%) |
| **Moderate to Severe MR** | 3 (1.0%) |
| **Abnormal Structure of AV & Thickened Heart Valves** | 2 (0.7%) |
| **Abnormal Structure of AV & Mitral Annular Calcification** | 2 (0.7%) |
| **AV Structural Abnormality & Abnormal Structure of AV & Mitral Annular Calcification & Thickened Heart Valves** | 2 (0.7%) |
| **AV Structural Abnormality & MV Structural Abnormality & Mitral Annular Calcification** | 2 (0.7%) |
| **AV Structural Abnormality & MV Structural Abnormality & Mitral Annular Calcification & Thickened Heart Valves** | 2 (0.7%) |
| **Moderate to Severe AR** | 2 (0.7%) |
| **TV Structural Abnormality** | 1 (0.3%) |
| **TV Structural Abnormality & MV Structural Abnormality & Mitral Annular Calcification & Thickened Heart Valves** | 1 (0.3%) |
| **AV Structural Abnormality & Thickened Heart Valves** | 1 (0.3%) |

| Condition | Count (%) |
|---|---|
| **Congenital Abnormalities & AV Structural Abnormality** | 1 (0.3%) |
| **Moderate to Severe TR** | 1 (0.3%) |
| **Moderate to Severe MR & MV Structural Abnormality** | 1 (0.3%) |
| **Moderate to Severe MR & MV Structural Abnormality & Mitral Annular Calcification & Thickened Heart Valves** | 1 (0.3%) |
| **Moderate to Severe MR & AV Structural Abnormality** | 1 (0.3%) |

AV: aortic valve, MV: mitral valve; TR: tricuspid regurgitation; TV: tricuspid valve
